# A human milk oligosaccharide alters the microbiome, circulating hormones, cytokines and metabolites in a randomized controlled trial of older individuals

**DOI:** 10.1101/2023.08.18.23294085

**Authors:** Matthew M. Carter, Diane Demis, Dalia Perelman, Michelle St. Onge, Christina Petlura, Kristen Cunanan, Kavita Mathi, Holden T. Maecker, Jo May Chow, Jennifer L. Robinson, Erica D. Sonnenburg, Rachael H. Buck, Christopher D. Gardner, Justin L. Sonnenburg

**Affiliations:** Department of Microbiology and Immunology, Stanford University School of Medicine, Stanford, CA, USA; Stanford Prevention Research Center, Stanford University School of Medicine, Stanford, CA, USA; Quantitative Sciences Unit, Stanford University School of Medicine, Stanford, CA, USA; Chan Zuckerberg Biohub, San Francisco, CA, USA; Human Immune Monitoring Center, Stanford University, School of Medicine, Stanford, CA 94304, United States of America; Abbott Nutrition, 3300 Stelzer Road, Columbus, OH 43219, USA; Center for Human Microbiome Studies, Stanford University School of Medicine, Stanford, CA, USA

## Abstract

Aging-related decline in immune function is associated with diseases like cancer, atherosclerosis, and neurodegenerative conditions. This study aimed to improve the aging gut microbiota and immune system by introducing a prebiotic oligosaccharide, 2-fucosyllactose (2’-FL), abundant in human breast milk with established health benefits in infants and animal models. 2’-FL was consumed at either of two doses versus placebo by 89 healthy older individuals (average age = 67.3 years) in a 6-week randomized controlled trial. Although the primary endpoint (significant change in the cytokine response score) was not met, consumers of the prebiotic experienced increased levels of *Bifidobacterium* in the gut microbiota, along with elevated serum levels of insulin, high-density lipoprotein (HDL) cholesterol, and fibroblast growth factor 21 (FGF21) hormone. Multi-omics analysis indicated a systemic response to 2’-FL, which could be detected in blood and urine, showcasing the potential of this prebiotic to provide diverse benefits to aging individuals.

## Introduction

Aging is associated with the broad decline of biological and cognitive function and the progression of diseases such as atherosclerosis and cancer ^1–3^. Central to this process of deterioration is immunosenescence, which is characterized by both chronic inflammation and hyporesponsiveness to stimuli ^4^. Combating the aging process will require the identification of therapeutic modalities that can counter immune system deterioration.

It is increasingly recognized that an important modifier of metabolic and immune system function is the trillions of microorganisms that comprise the human gut microbiota, or microbiome ^5^. Furthermore, microbiota-accessible carbohydrates (MACs), such as dietary fibers and resistant starches, have long been realized as potent modulators of the gut microbiota. Consumption of these complex carbohydrates by the microbiota not only results in the production of beneficial metabolites, but also may serve to properly tune the systemic immune system ^6^. Recent clinical trials investigating this premise have shown that dietary fiber supplementation in human cohorts results in heterogeneous, personalized responses that often but do not always result in a benefit to the host ^7–10^.

A type of microbiota-accessible carbohydrate that remains under-explored with regards to interventional studies in adult humans are the human milk oligosaccharides (HMOs). HMOs are the third most abundant constituent of human breast milk after lactose and lipids. They act as a key source of nutrients for key members of the developing infant gut microbiota, namely bifidobacteria ^11^. While HMOs cannot be directly metabolized by humans, they have recently been shown to have direct interactions with host cells independent of the gut microbiota ^12–14^. Pleiotropic benefits of HMOs for infants include fostering a healthy gut microbiota that protects against enteropathogens ^15,16^, training the immune system ^17^ and enhancing cognitive development ^18,19^. When combined with the potential for microbiota-dependent (prebiotic) and microbiota-independent (direct impact on host cells) effects, we hypothesized that HMOs could provide a benefit to adults. In sum, our study tests whether human biology and gut microbes possess extant pathways that would enable response to these molecules long after weaning. The capacity for HMOs to act as immune modulators in adults, especially aging adults who suffer from the sequelae of immunosenescence, requires detailed study.

## Results

### A randomized, placebo controlled clinical trial to study the effects of 2’-FL on the microbiota and host immune system

We performed the RAMP Study (Rejuvenating the Aging Microbiota with Prebiotics; ClinicalTrials.gov ID: NCT03690999) to investigate the effects of the HMO 2’-FL (**Figure 1A**) on the gut microbiota, immune system, and metabolism of elderly individuals. We recruited a cohort of 89 individuals (mean age = 67.4 years, minimum age = 60 years, maximum age = 83.9 years) and randomized them to one of three arms: a High Dose 2’-FL arm where subjects consumed 5g 2’-FL per day (n=29), a Low Dose 2’-FL arm where subjects consumed 1g 2’-FL per day (n=30), and a Placebo arm where subjects consumed a glucose product free of 2’-FL (n=30; **Figure 1B-C; Table S1; STAR methods**). The treatment period lasted for 6 weeks; 32 participants began the intervention after the start of the COVID-19 pandemic, resulting in modest deviations from the study protocol – primarily extending the duration of the protocol over the time that blood sampling facilities were locked down during the pandemic (these protocol deviations did not substantively affect the results of our study, see **Supplementary Note; Table S2**). We observed subjects starting 2 weeks before the intervention (Pre-baseline) as well as for 4 weeks after the intervention (Washout). Stool and blood samples were collected at Weeks −2, 0, 3, 6 and 10 (**Figure 1D**). We also collected anthropometrics and cognitive testing data at each of these time points. Urine was collected at Week 6.

**Figure 1.**
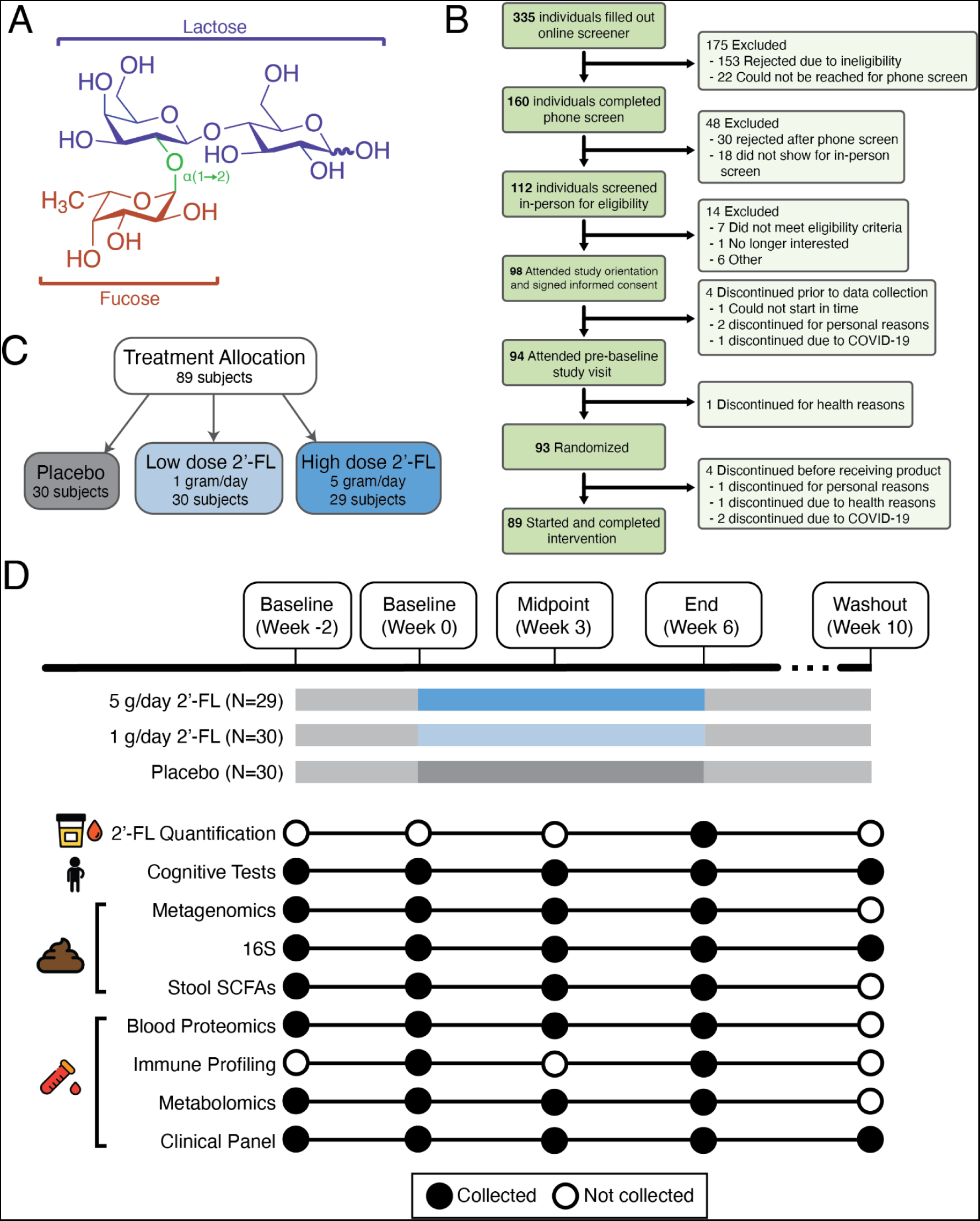
Design of a randomized controlled trial in healthy, elderly human subjects. (A) The chemical structure of 2’-FL. 2’-FL is composed of lactose (purple) covalently linked to a fucose (orange) via a glycosidic linkage (green). (B) A CONSORT Flow Diagram of the individuals recruited and screened for this study. (C) The treatment arm allocation of the 89 individuals who were randomized to a treatment arm and received the product. The three arms were: placebo, Low Dose 2’-FL (consuming 1 gram of 2’-FL per day; 0.5g AM and PM) and High Dose 2’-FL (consuming 5 grams of 2’-FL per day; 2.5g AM and PM). (D) Trial timeline (top) and sample collection schema (bottom). Each subject was tracked for a two-week of baseline period, supplementation began on Week 0 and continued for 6 weeks; participants were observed for a 4 week washout period after cessation of supplementation. Blood and stool were collected at each timepoint; urine was collected at the Week 6 timepoint. Subjects also took cognitive tests at each timepoint.

### Consumption of 2’-FL by adult subjects is well tolerated and leads to detectable 2’-FL in serum and plasma

Consumption of 2’-FL was well-tolerated and was not associated with any adverse events, such as gastrointestinal distress or nausea (**Figure S1**). To assess adherence to trial protocols and potential for systemic effects of 2’-FL we performed targeted mass spectrometry of 2’-FL in urine (n=66 samples) and plasma (n=89 samples) of each participant at the Week 6 timepoint (the lower number of urine samples available was due to COVID-related challenges of sample collection). Indeed, we detected 2’-FL in the plasma of 62% of the individuals in the High Dose 2’-FL arm and 13.3% of the individuals in the Low Dose 2’-FL arm compared to 0% of the individuals in the placebo arm. The High Dose 2’-FL group had significantly higher concentrations of 2’-FL compared to the placebo group for both urine and plasma (p=0.003 and p=9.6 x 10^-4^, respectively, Student’s T-test) (**Figure 2A-B**). This is, to our knowledge, the first time that an HMO has been measured in systemic circulation in non-lactating adult humans. Additionally, while systemic circulation of HMOs have been demonstrated in infants ^20^, lactating mothers ^21^ and animal models ^22^, the mechanism of translocation from the lumen of the gastrointestinal tract to the bloodstream is unclear. *In vitro* data suggest paracellular transport and/or receptor-mediated transcytosis may be involved ^23^.

**Figure 2.**
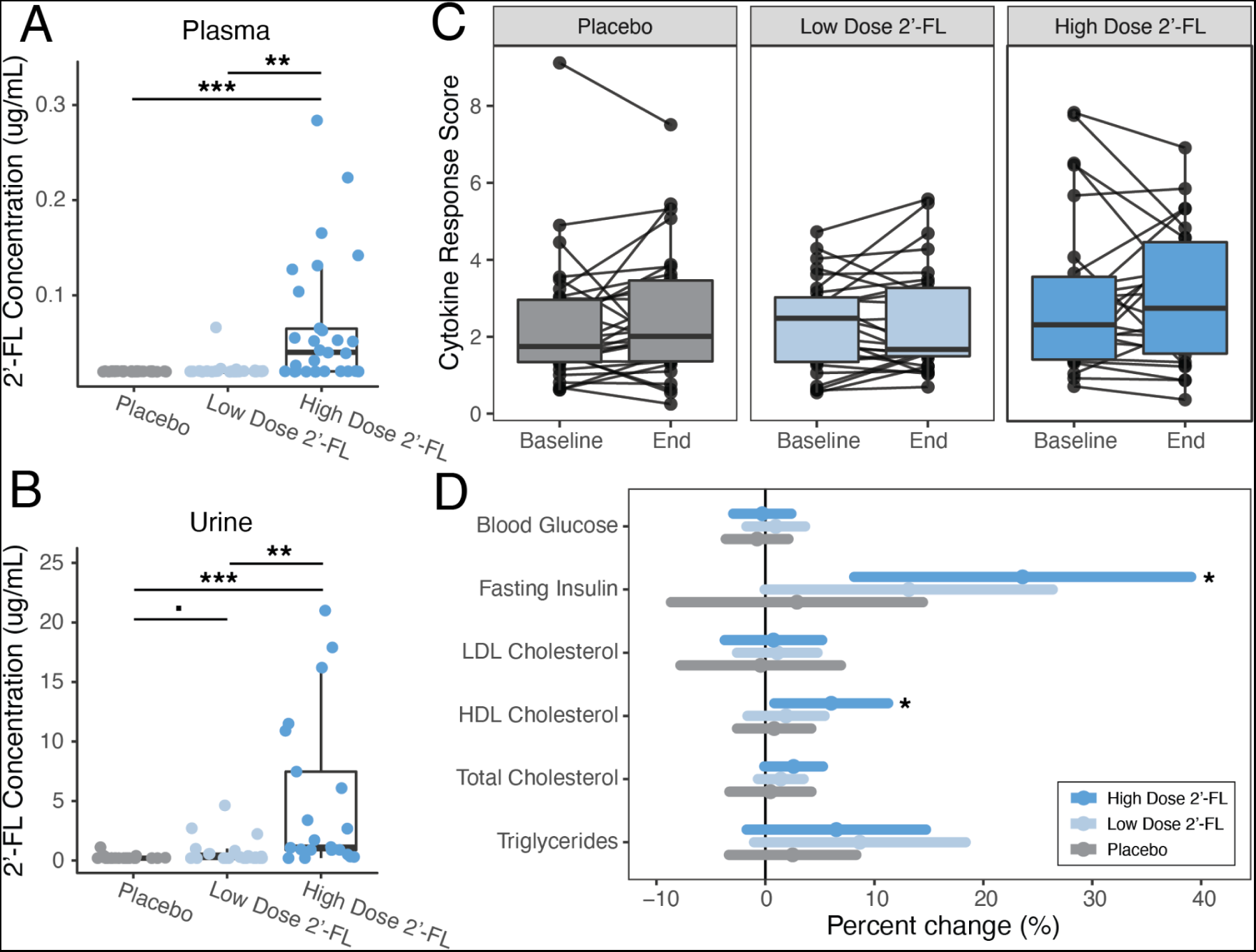
2’-FL is detectable in plasma and urine of the High Dose 2’-FL group and results in significant increases in fasting insulin and HDL cholesterol, although it doesn’t affect Cytokine Response Score. (A) 2’-FL is detected at significantly greater levels in plasma at the Week 6 timepoint in the High Dose 2’-FL group compared to both the Low Dose 2’-FL group (p = 0.001, Student’s T-test) and the Placebo group (p = 9.6 x 10^-4^, Student’s T-test). (B) 2’-FL is detected at significantly greater levels in urine at the Week 6 timepoint in the High Dose 2’-FL group compared to both the Low Dose 2’-FL group (p = 0.006, Student’s T-test) and the Placebo group (p = 0.003, Student’s T-test). The Low Dose 2’-FL group trended towards higher concentrations of 2’-FL compared to the Placebo group (p = 0.069, Student’s T-test). (C) Cytokine Response Score does not change significantly from Baseline (Week 0) to End of Intervention (Week 6) for any of the three treatment groups. (D) Percent change from Baseline (Week 0) to End of Intervention (Week 6) for metabolic markers. Fasting insulin and HDL cholesterol increased significantly from Week 0 to Week 6 in the High Dose 2’-FL group but not the Low Dose 2’-FL group or the Placebo group. Dots are the mean change from Baseline (Week 0) to End of Intervention (Week 6) and lines are 95% confidence intervals of the means.

### Consumption of 2’-FL had no effect on Cytokine Response Score but is associated with increases in HDL cholesterol and fasting insulin

The pre-registered primary outcome of the study was change in the Cytokine Response Score (CRS) from baseline to Week 6. The CRS was defined previously as a set of features within immune profiles that discriminated younger from older individuals ^4^; these features indicate hyporesponsiveness in peripheral blood mononuclear cells (PBMCs) of older individuals, and we reasoned that change in CRS may indicate a reversal of inflammation associated with aging. We did not observe significant changes in CRS between treatment groups or from baseline to end of intervention within any of the three groups (**Figure 2C**).

We used a standard clinical panel to measure the effect of 2’-FL on host metabolic status. We found that the High Dose 2’-FL group, but not the Placebo or Low Dose groups, had significantly increased high-density lipoprotein cholesterol at End of intervention (Week 6) compared to Baseline (Week 0) (95% confidence interval: 0.8% − 11.3%) while low-density lipoprotein (LDL) cholesterol was unchanged (95% confidence interval: −3.7% decrease − 5.2% increase). We also found that fasting insulin increased significantly and specifically in the High Dose 2’-FL group (95% confidence interval: 8.1% − 39.1% increase) (**Figure 2D**). While higher fasting insulin is sometimes associated with insulin insensitivity ^24^ no subjects’ fasting insulin values exceeded reference ranges during Week 6 for the High Dose 2’-FL group (**Figure S2**).

### Bifidobacterium blooms in response to consumption of 2’-FL

We performed 16S rRNA sequencing on each stool sample collected in order to assess compositional changes in the microbiota during the course of the trial (**Table S3**). We did not find statistically significant changes in Shannon diversity over the course of the intervention (**Figure 3A**). A beta diversity analysis revealed a significant association between gut microbiota composition with treatment arm at Week 3 (P=0.03, Adonis test, **Figure 3B**) but not Week 0 (P=0.99, Adonis test) or Week 6 (P=0.35, Adonis test). Comparing individual treatment groups at Week 3 revealed that the High Dose 2’-FL group was significantly different from the Placebo group (P=0.006, Adonis test). We performed metagenomic sequencing to determine if microbiota functional capacity was affected by 2’-FL supplementation and found that a small but statistically significant portion of variation in KEGG pathway beta diversity was explained by the treatment arm (P<0.001, Adonis test; **Figure S3A**).

**Figure 3.**
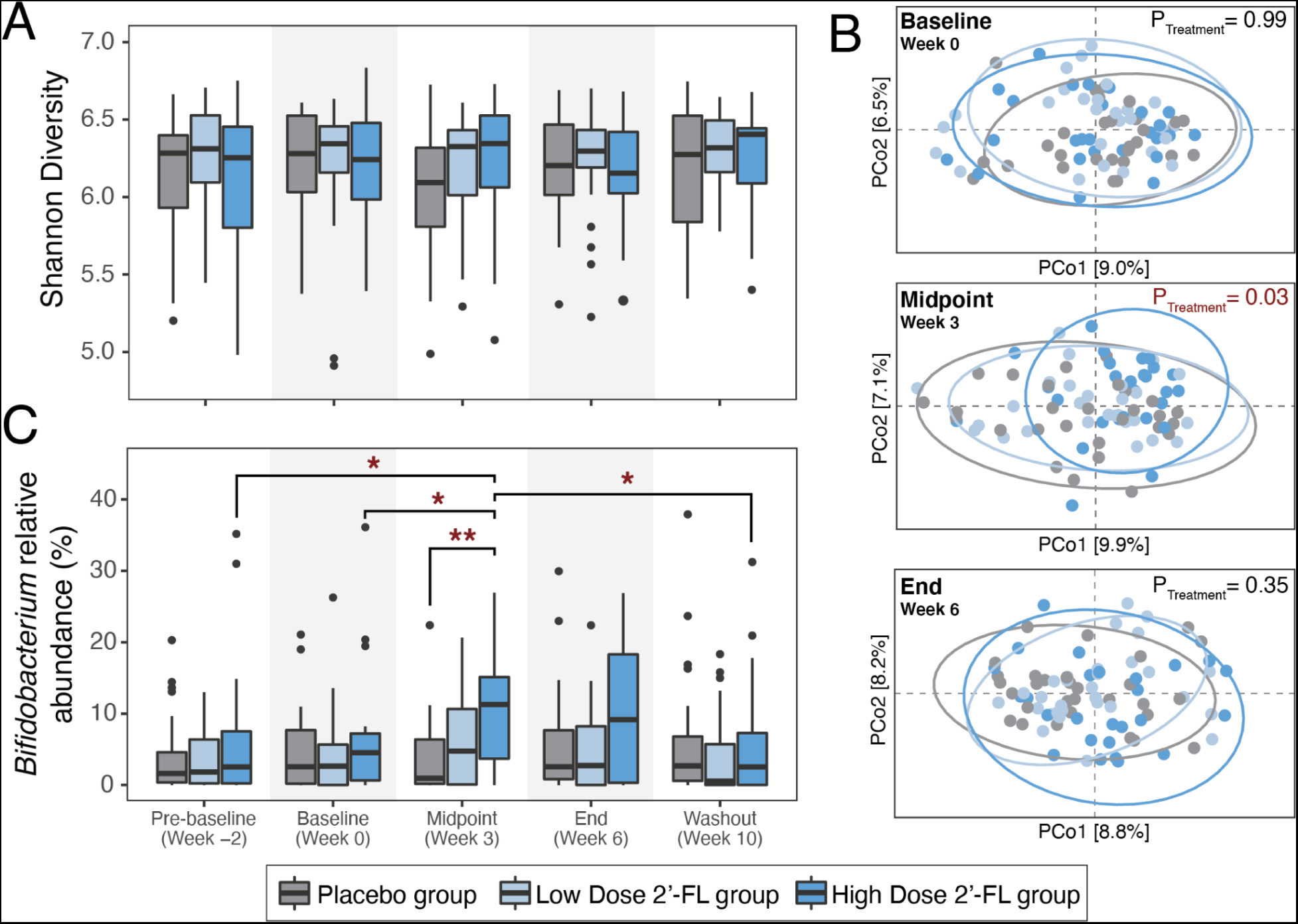
2′-FL induces a bloom of *Bifidobacterium* that is most prominent in Week 3 and which subsides to baseline levels during the washout period. (A) Shannon (alpha) diversity of each treatment group at each timepoint during the intervention. (B) Beta diversity of each treatment group at Baseline (Week 0), Midpoint (Week 3) and End of intervention (Week 6), depicted via principal coordinate analyses (PCoA). Treatment group explained a significant portion of variation at the Midpoint (Week 3) timepoint only (P=0.03, Adonis test). (C) Relative abundance of the genus *Bifidobacterium* for each treatment group at each timepoint during the intervention. The High Dose 2’-FL group had higher *Bifidobacterium* relative abundance at Midpoint (Week 3) compared to the Placebo group (P=0.0016, Wilcoxon rank-sum test) and also compared to the High Dose 2’-FL group at the Pre-Baseline Week −2 (P=0.01, Wilcoxon rank-sum test), Baseline Week 0 (P=0.037, Wilcoxon rank-sum test) and Washout Week 10 (P=0.037, Wilcoxon rank-sum test) timepoints.

We hypothesized that the taxa responsible for driving this change in composition was the genus *Bifidobacterium*, whose member species are known to be specialist degraders of HMOs in the developing infant microbiota ^25,26^, but also inhabit the adult GI tract. Indeed, relative abundance of *Bifidobacterium* was significantly higher in the High Dose 2’-FL group (mean relative abundance = 9.7%, standard error (s.e.) = 1.4%) compared to the Placebo group (mean relative abundance = 3.5%, s.e. = 0.96%) at the study midpoint (Week 3) (**Figure 3C**). The relative abundance of *Bifidobacterium* in the Low Dose 2’-FL group was not significantly different from the Placebo or the High Dose 2’-FL groups at Week 3 but was at an intermediate abundance between both groups (mean relative abundance = 6.0%, s.e. = 1.3%). *Bifidobacterium* relative abundance was also significantly higher for the High Dose 2’-FL group at the midpoint of the study (Week 3) compared to the same group at baseline (Week −2 and Week 0) and at the end of the study (Week 10) (P=0.015, P=0.037 and P=0.035, respectively, paired Wilcoxon rank-sum tests).

Analysis of the relative abundance of individual species of *Bifidobacterium* which we could detect (*B. adolescentis, B. catenulatum, and B. breve*) over time across groups revealed one species that followed a similar trend as the genus: *Bifidobacterium adolescentis*. *B. adolescentis* had greater relative abundance at the Week 3 timepoint in the High Dose 2’-FL group compared to the Placebo groups (P=0.001, Wilcoxon rank-sum test, **Figure S3B**). The High Dose 2’-FL group also had greater relative abundance of *B. catenulatum* compared to the Placebo group, but this was not statistically significant (P=0.056, Wilcoxon rank-sum test). Relative abundance of *B. breve* remained unchanged over time and between study arms. Analysis of all genera in the gut microbiota revealed that no other taxa had significantly increased or decreased relative abundance during the intervention. The data, taken together, demonstrate the strongest microbiome effect in the Week 3 time point which subsided to baseline levels during the post-intervention washout period, a resilience-effect consistent with other studies examining dietary microbiota perturbations ^27,28^.

*Bifidobacterium spp.* are known to ferment HMOs to produce the short-chain fatty acid (SCFA) acetate and also provide lactate to other butyrate-producing bacteria ^29^. We next sought to determine whether consumption of 2’-FL led to increases in the production of SCFAs by the microbiota. We performed targeted metabolomics on stool samples collected from each subject at Week 0 and Week 6 to determine the stool concentrations of 9 SCFAs (acetate, propionate, butyrate, 2-methylpropionate, pentanoate, 3-methylbutyric, 2-methylbutyric, hexanoate, and 4-methylpentanoate). We specified a linear mixed effects model (LMM) for each of these SCFAs based on treatment arm and time point and did not observe any statistically significant effects. We next correlated stool SCFA concentrations with the relative abundance of each genera detected in these stool samples. We found that *Akkermansia* was negatively associated with acetate and propionate (P=0.0056 and P=0.0073, respectively, LMM, Benjamini-Hochberg correction) and *Dorea* was negatively associated with 2-methylbutyric, 3-methylbutyric and 2-methylpropionate (P=0.032, P=0.048 and P=0.037, respectively, LMM, Benjamini-Hochberg correction). We also found that *Dialister* was positively associated with acetate, propionate and butyrate (P=1.72 x 10^-5^, P=3.64 x 10^-4^ and P=0.023, respectively, LMM, Benjamini-Hochberg correction) and that *Holdemanella* was positively associated with acetate and propionate (P=0.0066 and P=0.0015, respectively, LMM, Benjamini-Hochberg correction) (**Figure S4A**). We next specified linear mixed effects models to determine if components of diet (dietary fiber, total carbohydrates, total protein or total energy) correlated with stool SCFA. While dietary fiber did not demonstrate statistically significant correlations with SCFAs, we found that total caloric intake was significantly associated with acetate, propionate and hexanoate (P=0.045, P=0.030 and P=0.032, respectively, LMM, Benjamini-Hochberg correction; **Figure S4B**).

### Metabolites and circulating inflammatory proteins are altered in 2’-FL consuming individuals

We next sought to understand how 2’-FL consumption might affect the host metabolic and immune status. To this end, we analyzed serum samples from each subject with both untargeted metabolomics on 127 circulating metabolites (**Table S4**) and targeted proteomics on 92 circulating cytokines, chemokines and inflammation-associated proteins (**Table S5**). We specified linear mixed effects models for each metabolite and immune proteins in order to discover metabolites and proteins that are enriched or depleted during the intervention when comparing High Dose 2’-FL vs. Placebo, Low Dose 2’-FL vs. Placebo, and High Dose 2’-FL vs. Low Dose 2’-FL.

We found that octanoylcarnitine was depleted in the High Dose 2’-FL group compared to both the Low Dose 2’-FL group (P=0.004, LMM, Benjamini-Hochberg correction, **Figure 4A**, **Table S6**) and the Placebo group (P=7.3×10^-4^, LMM, Benjamini-Hochberg corrected). Octanoylcarnitine is a medium-chain fatty acid covalently linked to carnitine which undergoes fatty acid oxidation in the mitochondria. We also found that uracil and glutamate were enriched in the High Dose 2’-FL group compared to the Low Dose 2’-FL group (P= 0.017 and P= 0.017, respectively, LMM, Benjamini-Hochberg correction). There were no metabolites that were differentially enriched when comparing the Low Dose 2’-FL group to the Placebo group.

**Figure 4.**
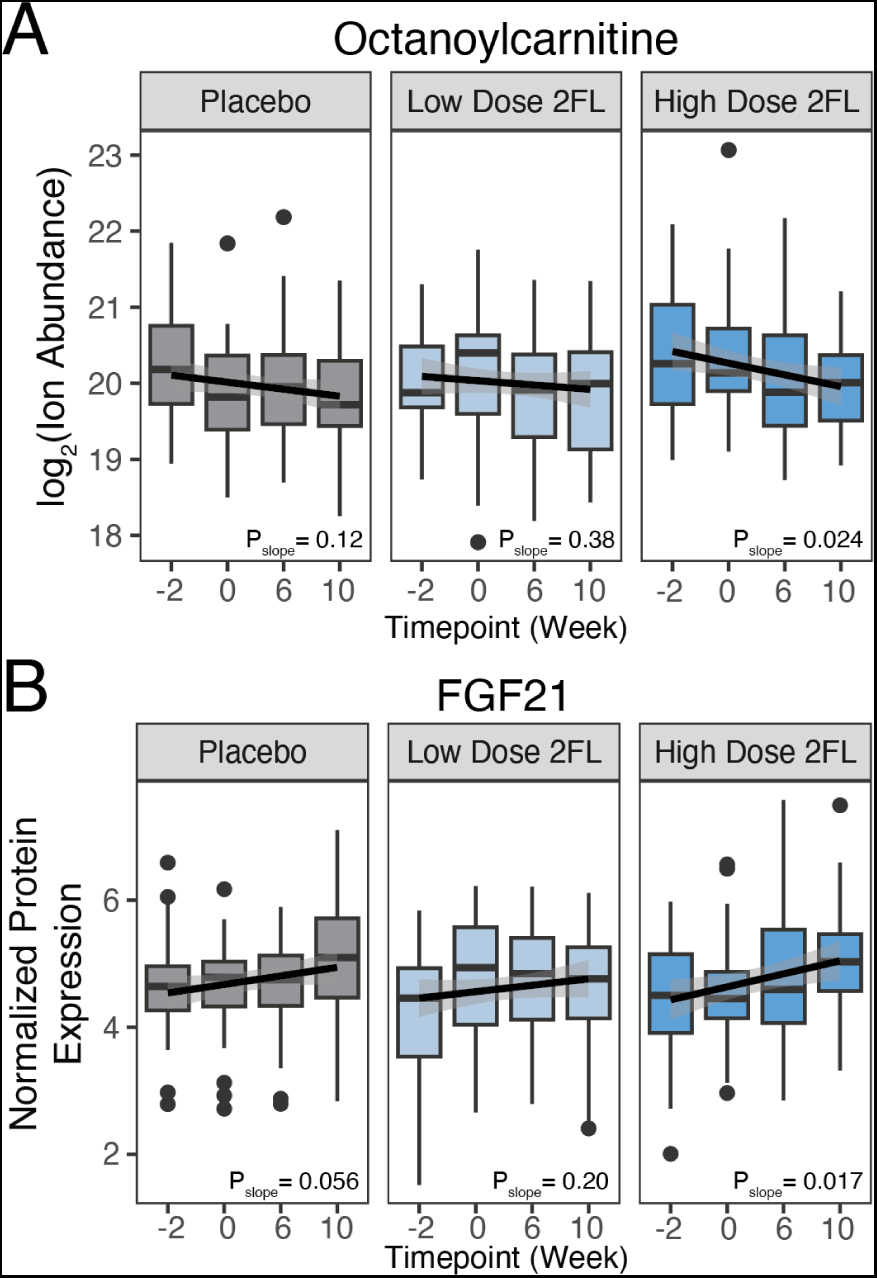
The metabolite octanoylcarnitine and the hormone FGF21 are altered in the serum of the High Dose 2’-FL group compared to the Placebo and Low Dose 2’-FL groups. (A) Boxplots showing the log2 abundance of octanoylcarnitine from untargeted metabolomics analysis of serum samples. Three panels from left to right show metabolite abundance for Placebo group, Low Dose 2’-FL group and High Dose 2’-FL group, respectively, over time. Solid black lines are trendlines for the linear regression of count versus time for each treatment group. P-values are the significance of the slopes of regressions. (B) Boxplots showing the normalized protein expression for FGF21 from Olink proteomics analysis of serum samples. Three panels from left to right show protein abundance for Placebo group, Low Dose 2’-FL group and High Dose 2’-FL, respectively, group over time. Solid black lines are trendlines for the linear regression of count versus time for each treatment group. P-values are the significance of the slopes of regressions.

With regards to circulating proteins, we found that fibroblast growth factor 21 (FGF21) was significantly elevated in the High Dose 2’-FL group vs. the Placebo group (P=0.004, LMM, Benjamini-Hochberg correction, **Figure 4B**) and also elevated in the High Dose 2’-FL group compared to the Low Dose 2’-FL group (P=0.003, LMM, Benjamini-Hochberg correction). FGF21 is a liver secreted hormone that regulates lipid metabolism ^30^. In human clinical trials, FGF21 analogues have been shown to increase HDL cholesterol levels while down-regulating lipolysis ^31,32^, consistent with our finding of increased HDL cholesterol (**Figure 2B**) and decreased serum octanoylcarnitine (**Figure 4A**) in the High Dose 2’-FL group. We also found that CD40 and the sulfotransferase ST1A1 were upregulated in the High Dose 2’-FL group compared to the Low Dose 2’-FL group (P=0.041 and P=0.049, respectively, LMM, Benjamini-Hochberg correction).

### Increased *Bifidobacterium* abundance corresponds to more Bifidobacterium at baseline and circulating 2’-FL at end of intervention

We observed a heterogeneous response to 2’-FL consumption with regards to changes in *Bifidobacterium* relative abundance during the intervention. This heterogeneity may be due to i) lack of *Bifidobacterium* in the gut microbiota at baseline, ii) lack of *Bifidobacterium* that is capable of degrading 2’-FL, or iii) other ecological factors. To provide insight into whether the metabolomic and proteomic changes observed in the 2’-FL treatment groups may depend on a bloom of *Bifidobacterium* in the gut microbiota, we subsetted participants from the High Dose 2’-FL and Low Dose 2’-FL groups based on their change in *Bifidobacterium* relative abundance from Week 0 to Week 3. Participants above the median *Bifidobacterium* change were categorized as “responders” (n=30 participants) and those below the median were categorized as “nonresponders” (n=29 participants) (**Figure 5A**). Of the 30 responders, 18 were in the High Dose 2’-FL group and 12 were in the Low Dose 2’-FL group. While responders had greater *Bifidobacterium* relative abundance at Week 0 (baseline) on average compared to nonresponders, this difference was not statistically significant (P=0.055, Wilcoxon rank-sum test, **Figure 5B**). Interestingly, we found that responders had higher levels of 2’-FL in plasma at Week 6 compared to nonresponders (P=0.038, Wilcoxon rank-sum test, **Figure 5C**).

**Figure 5.**
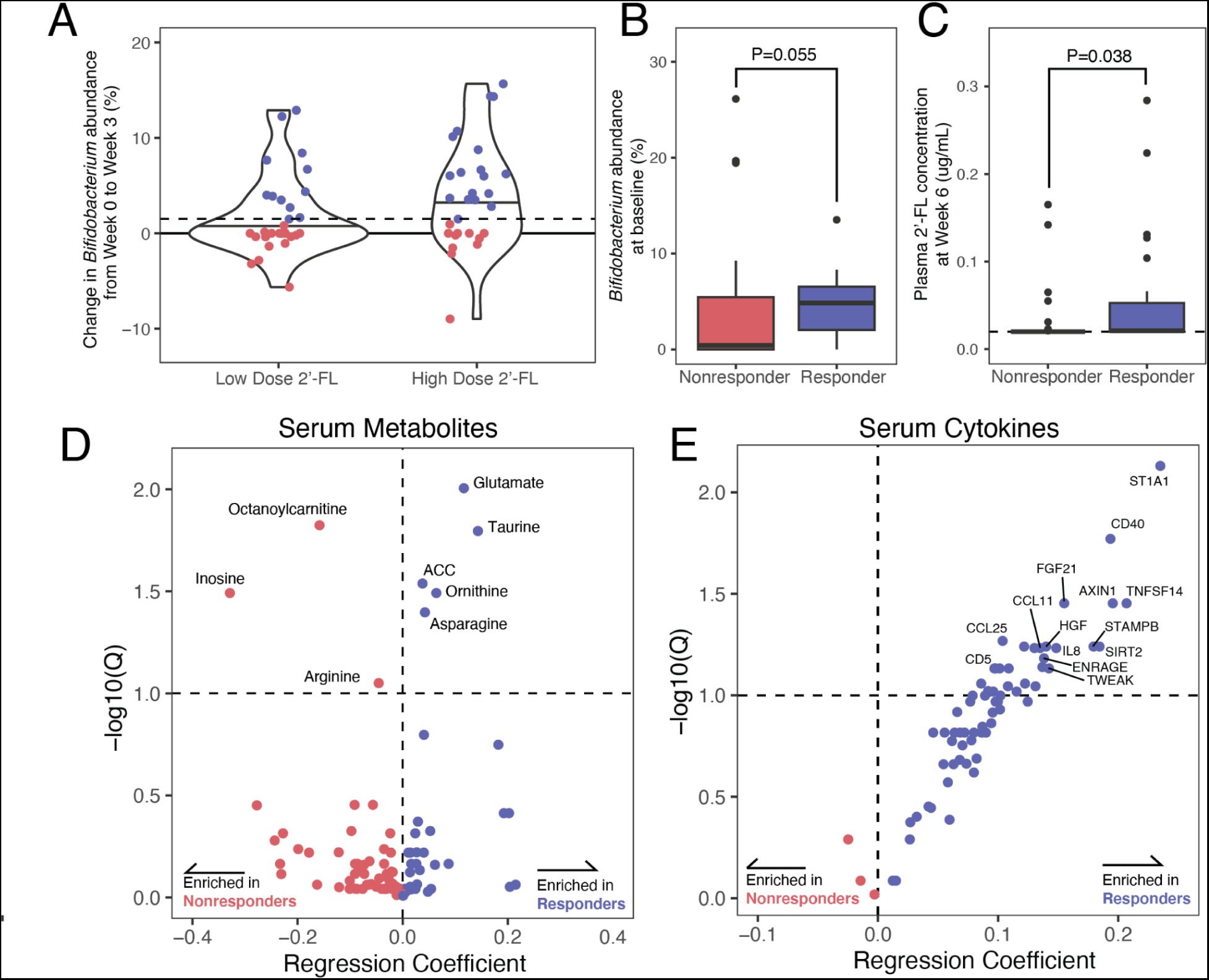
Sub-setting subjects into responders and nonresponders based on change in *Bifidobacterium* abundance reveals widespread changes to levels of circulating cytokines. (A) Subsetting of subjects into responders and nonresponders based on change in *Bifidobacterium* relative abundance between Baseline (Week 0) and Midpoint (Week 3). Participants were classified as responders if they were in the top tertile of *Bifidobacterium* increase from Week 0 to Week 3. (B) *Bifidobacterium* relative abundance at Baseline (Week 0) for responders and nonresponders is not significantly different (P=0.055, Wilcoxon rank-sum test). (C) Plasma concentrations of 2’-FL at End of intervention (Week 6) for responders and nonresponders. Responders have significantly greater plasma concentrations of 2’-FL (P=0.038, Wilcoxon rank-sum test). (D) Volcano plot of untargeted serum metabolomics showing the metabolites enriched in responders and nonresponders. P-values were false-discovery rate corrected with the Benjamini-Hochberg method. Horizontal dashed line represents a q-value threshold of 0.1. (ACC = 1-aminocyclopropane-1-carboxylic acid). (E) Volcano plot of targeted serum proteomics showing the proteins enriched in responders and nonresponders. P-values were false-discovery rates corrected with the Benjamini-Hochberg method. Horizontal dashed line represents a q-value threshold of 0.1.

### Serum metabolites and cytokines are enriched in responders

Next, we sought to determine whether any physiological consequences corresponded to the observed bloom in *Bifidobacterium* abundance. We re-analyzed the untargeted metabolomics data, this time comparing responders versus nonresponders, and discovered 5 metabolites enriched in responders (glutamate, taurine, ornithine, asparagine and 1-aminocyclopropane-1-carboxylic acid) and 3 metabolites enriched in nonresponders (octanoylcarnitine, inosine and arginine; **Figure 5D**). Pathway analysis of these 8 metabolites reveals a significant impact on the arginine biosynthesis pathway (P = 9.71×10^-4^, Benjamini-Hochberg corrected).

We found that responders showed significant enrichment for 24 circulating cytokines compared to nonresponders (**Figure 5E**). Network analysis of these cytokines with STRING revealed a local clustering coefficient 0.591 (P<1×10^-16^). Gene Ontology pathway analysis revealed significant pathway enrichment of “cytokine-mediated signaling pathway”, “cellular response to organic substance”, and “neutrophil chemotaxis” (P=1.00×10^-13^, 7.36×10^-10^, 8.38×10^-9^, respectively, Benjamini-Hochberg FDR correction). Notably, one significantly up-regulated protein, sirtuin 2 (SIRT2), has been strongly implicated in aging and longevity ^33^.

In order to ensure that we were not arbitrarily subsetting our data in a way that inflated our false discovery rate, we simulated the responder/nonresponder enrichment analysis 500 times, randomly permuting the responder and nonresponder labels each time. For the metabolomics analysis the “true” responder/nonresponder labels yielded more significant features than >94% of simulated analyses and for the proteomics analysis the “true” responder/nonresponder labels yielded more significant features than >84% of simulated analyses (**Figure S5**). This sensitivity analysis underscores the utility of our responder subsetting scheme, showing that *Bifidobacterium*-based subsetting of participants coincides with a broader biological impact of the intervention.

### Multi-omics associations between serum metabolomics and cytokines

In addition to understanding the impact of a dietary intervention on human subjects, multi-omics studies such as this one can be used to discover novel interactions between different facets of host biology. To this end, we next sought to understand how changes in circulating levels of serum metabolites and serum cytokines might influence one another. We calculated the baseline-to-end fold change of each metabolite and cytokine for each participant and performed the all-versus-all correlations across the combined data sets (**Figure 6A**). We discovered extensive, significant associations both within and between these data sets. Eleven metabolites and 18 serum proteins were implicated in statistically significant cross-data set correlations after multiple hypothesis testing correction (**Figure 6B**). Six of these 11 metabolites are purines (or purine derivatives) and 4 of these 11 metabolites are amino acids (or amino acids derivatives). Notably, 5 of these metabolites (asparagine, inosine, glutamate, taurine, and ornithine) and 10 of these cytokines (AXIN1, CD40, ENRAGE, HGF, OSM, SIRT2, ST1A1, STAMPB, TGF-alpha, and TNFSF14) were also significantly enriched in *Bifidobacterium* responders or nonresponders, indicating a broad rewiring of the host metabolome and immuno-proteome in response to the 2’-FL-mediated *Bifidobacterium* bloom.

**Figure 6.**
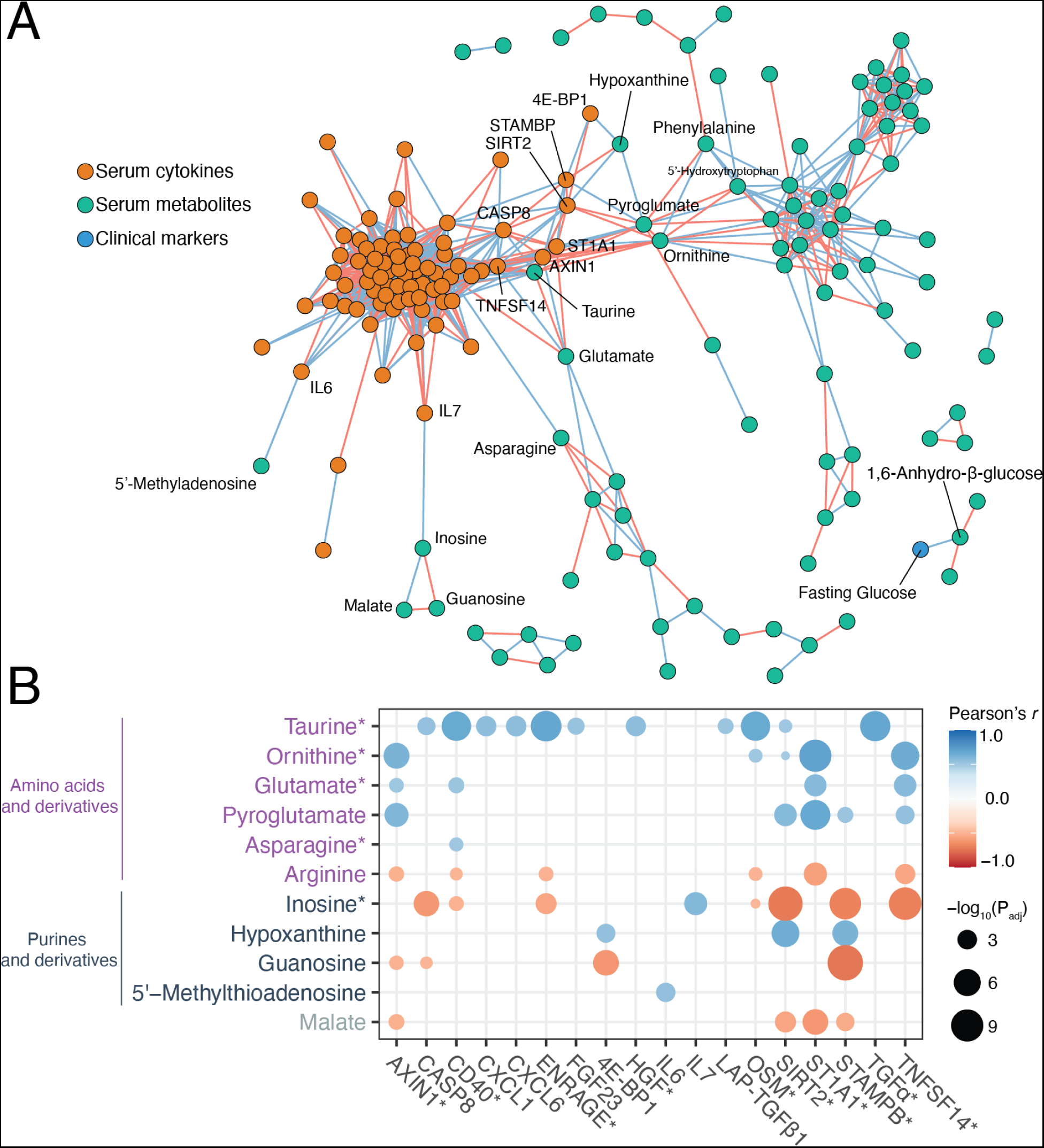
Multi-omic associations between serum metabolomics and serum cytokine proteomics. (A) A correlation network between serum metabolomics (green nodes), serum cytokine proteomics (orange nodes) and clinical markers (blue nodes). All-versus-all correlation network was based on centered and scaled baseline-to-end changes in individual metabolite/protein levels across all participants. Blue edges indicate positive correlation, red edges indicate negative correlation. (B) Significant pairwise correlations between serum metabolites and serum cytokines using Pearson correlation (p-value correction with Benjamini-Hochberg method, corrected p-values ≤ 0.05). Larger circles indicate more significant corrected p-values. Color map indicates the strength of the Pearson correlation with blue indicating positive correlation and red indicating negative correlation. Metabolites and cytokines with asterisks were significantly associated with responder status.

### Responders perform better on assessments of cognitive abilities

During the course of the trial participants took two forms of cognitive assessments from the Cambridge Neuropsychological Test Automated Battery (CANTAB) of computer-based tests ^34^: the Rapid Visual Information Processing (RVP) test, which assesses sustained attention and the Paired Associates Learning (PAL) test, a measure of visual memory and learning. For both tests, there was no significant difference in performance between treatment groups (P=0.81 and P=0.94 for PAL and RVP, respectively, analysis of covariance analysis; **Figures S6A-B**). However, *Bifidobacterium* responders performed comparatively better than nonresponders on the PAL test (P=0.016 analysis of covariance, **Figure S6C**). There were no significant differences between responders and nonresponders for the RVP test (P=0.081, analysis of covariance; **Figure S6D**). These results, while speculative, are in line with research in infants and animal models regarding the effect of 2’-FL on cognition and memory ^18,19,35^ and suggest the need for future studies designed to test cognitive performance explicitly.

## Discussion

The aging immune system is characterized by the impaired response to stimuli as a result of heightened baseline inflammation ^4^. This immune dysregulation is implicated in numerous non-communicable pathologies such as cancer, atherosclerosis, and neurodegeneration ^1,2^. Our work demonstrates that dosing older adult humans with the human milk oligosaccharide 2′-fucosyllactose (2′-FL) results in a targeted modification of the microbiota, namely a bloom of *Bifidobacterium* in the High Dose 2′-FL group. This bloom of *Bifidobacterium* was accompanied by widespread metabolic and immune alteration. Given that we show evidence of 2’-FL entering systemic circulation (**Figure 2A-B**) and that past work shows direct effects of 2’-FL on host cells ^12,13^, future work will be needed to determine the extent to which these HMO-mediated effects are dependent upon activity of the microbiota.

We observed two significant changes in the clinical panels of participants in the High Dose 2’-FL group: significant increases in fasting insulin and HDL cholesterol. Previous work has shown that *Bifidobacterium spp.* can induce glucagon-like peptide-1 (GLP-1) secretion in the gastrointestinal tract ^36^. GLP-1 in turn, stimulates the secretion of insulin ^37^. *Bifidobacterium spp*. are also known to affect cholesterol and lipid homeostasis through the activity of bile salt hydrolases. These results suggest that the increase in *Bifidobacterium* that we observed in the High Dose 2’-FL group may be responsible for the changes to circulating insulin and HDL cholesterol. Directly measuring bacteria, metabolites and hormones in the gastrointestinal tract *in situ* is challenging in human studies and future studies in animal models will help to uncover host-microbe interactions occurring in this part of the gastrointestinal tract.

*Bifidobacterium spp.* are widely used as probiotic agents and are associated with a panoply of host benefits ^38^ which makes their enrichment in the gut microbiota via HMOs an attractive therapeutic target. Oral delivery of probiotics has demonstrated mixed but promising clinical value ^39–41^. Furthermore, many probiotic formulations may fail to properly engraft in the host microbiota. Recent studies have shown improved engraftment of probiotic strains when they are delivered as “synbiotics” with prebiotic compounds ^42,43^. We demonstrate a modulation of *Bifidobacterium* abundance in human subjects via consumption of an HMO without the use of a probiotic supplement. A drawback of this HMO-only prebiotic approach is that metabolic and immunological benefits seem to be individualized. Studies of probiotic engraftment have shown responses are contingent upon the baseline composition of an individual’s microbiota ^44^. Additionally, this 2′-FL-mediated modulation of *Bifidobacterium* was reversible: relative abundance subsided to baseline levels after cessation of prebiotic consumption. Similar findings have been reported with other prebiotics ^42,45^. These personalized kinetics of “recovery” from dietary interventions with prebiotics are analogous to the microbiota resilience that follows cessation of a variety of perturbations such as seminal studies of recovery from antibiotics ^46^. In this study, the high dose of 5g/day was well-tolerated and resulted in prominent changes to the gut microbiota as well as to the host metabolome and proteome. One recent study showed well-tolerated consumption of up to 20g/day 2′-FL in younger adults ^47^ and another showed 18 g/day of a mixture of HMOs derived directly from breastmilk^42^. Longer term intervention studies will be needed to understand the nature of resiliency in the gut microbiota during and after a dietary intervention. Future studies will also be needed to understand proper dosing regimens for HMOs and whether administration of live bifidobacteria or other bacteria are required to achieve specific individualized versus generalized responses in adult subjects.

## Data Availability

The authors declare that the data supporting the findings of this study are available within the paper and its supplementary information files. Metagenomic and 16S reads are being submitted to the short read archive (SRA) under the BioProject PRJNA996611 and this manuscript will be updated with additional accessions when the submission is complete. Code and analysis files for this project can be found at https://github.com/SonnenburgLab/RAMP_study_analysis.

## Acknowledgements

We wish to thank the participants for their engagement and effort to enable this study, Hannah Wastyk for providing guidance and mentorship, the University of Minnesota Genomics Center for 16S sequencing, Tala Khosroheidari and William Beckwith at Olink proteomics, the Human Immune Monitoring Core for staining and running phosphoflow and O-link assays, the Chan-Zuckerberg Biohub for metagenomic sequencing, Metabolon for providing targeted mass spectrometry analysis, Yuqin Dai and the Metabolomics Knowledge Center for supporting mass spectrometry analysis, Will Van Treuren for aiding with metabolomics method development and the Stanford Clinical and Translational Research Unit. This work was funded by Abbott Nutrition. M.M.C. was supported by a Stanford Graduate Fellowship. J.L.S. is a Chan-Zuckerberg Biohub investigator.

## Author contributions

M.M.C., E.D.S., C.D.G., and J.L.S. performed the experiments and designed and performed data analysis. D.D. and D.P. provided patient interaction, dietary counseling, and analysis. M.S.O., A.W., and M.M.C. performed stool aliquoting, metagenomic sequencing and analysis. K.M., Y.R.H., and H.T.M. performed immune profiling assays. K.C. aided with statistical analyses. J.L.R. and D.D. oversaw patient recruitment and study design implementation. M.M.C., C.D.G., J.L.S., J.M.C., and R.H.B. conceived of the study and wrote the manuscript.

## Supplemental Information for

### Supplementary Note

#### Description of COVID-19-related protocol deviations

Between mid-March to the end of May 2020, participants were not able to come in for a blood draw as residents of California and the Bay Area were mandated to shelter-in-place. Furthermore, the Clinical Translational Research Unit (CTRU) was unexpectedly closed down and open only to essential research appointments, of which the RAMP Study was not included.

Fifteen participants were in the intervention phase of the study when COVID-19-related shelter orders and facility shutdowns occurred. Rather than drop the participants, we mailed the participants additional bottles of their assigned supplement (either the placebo supplement or the 2’-FL-containing supplement). The table below lists the participant IDs of each affected participant as well as the number of additional days that they consumed the supplement. Of these 15 participants affected by the shutdown, 5 were in the Placebo arm, 4 were in the Low Dose 2’-FL arm and 6 were in the High Dose 2’-FL arm.

#### Sensitivity analysis of COVID-19 related protocol deviations

In order to understand the effect of these deviations on our results, we performed a sensitivity analysis on the results from **Figure 2** (clinical findings and primary outcome) and **Figure 3** (microbiome results).

In the two figures below, we re-analyze the results from **Figure 2** and **Figure 3** by subsetting individuals into two groups: “No COVID-19 Deviations” if they stayed on the pre-specified protocol and “Experienced COVID-19 Deviations” if they deviated from the pre-specified protocol by continuing to take their assigned supplement for the duration listed in **Table S2**. We perform the same statistical analyses as on the full data set and report the findings in **Figure D1** (primary outcome and clinical data) and **Figure D2** (microbiome data).

We found that there were no qualitative differences in outcomes between the COVID-19 protocol deviation group and the non-deviation group with regards to 2’-FL levels in plasma and the primary outcome (Cytokine Response Score). We found that there were minor differences in the clinical outcomes for the COVID-19 protocol deviation group, but these results are hard to interpret given the small sample size of the deviation group (particularly the Low Dose 2-FL subset of this group, which was n=4). With regards to microbiome data, we found no qualitative differences between the COVID-19 protocol deviation group and the non-deviation group with regards to Shannon diversity and *Bifidobacterium* abundance. Both groups showed no significant differences in Shannon diversity over time or between groups, and both groups had statistically significant increases in *Bifidobacterium* at the Week 3 time point compared to Placebo and that the increases subsided back to baseline during the washout period.

**Figure D1.**
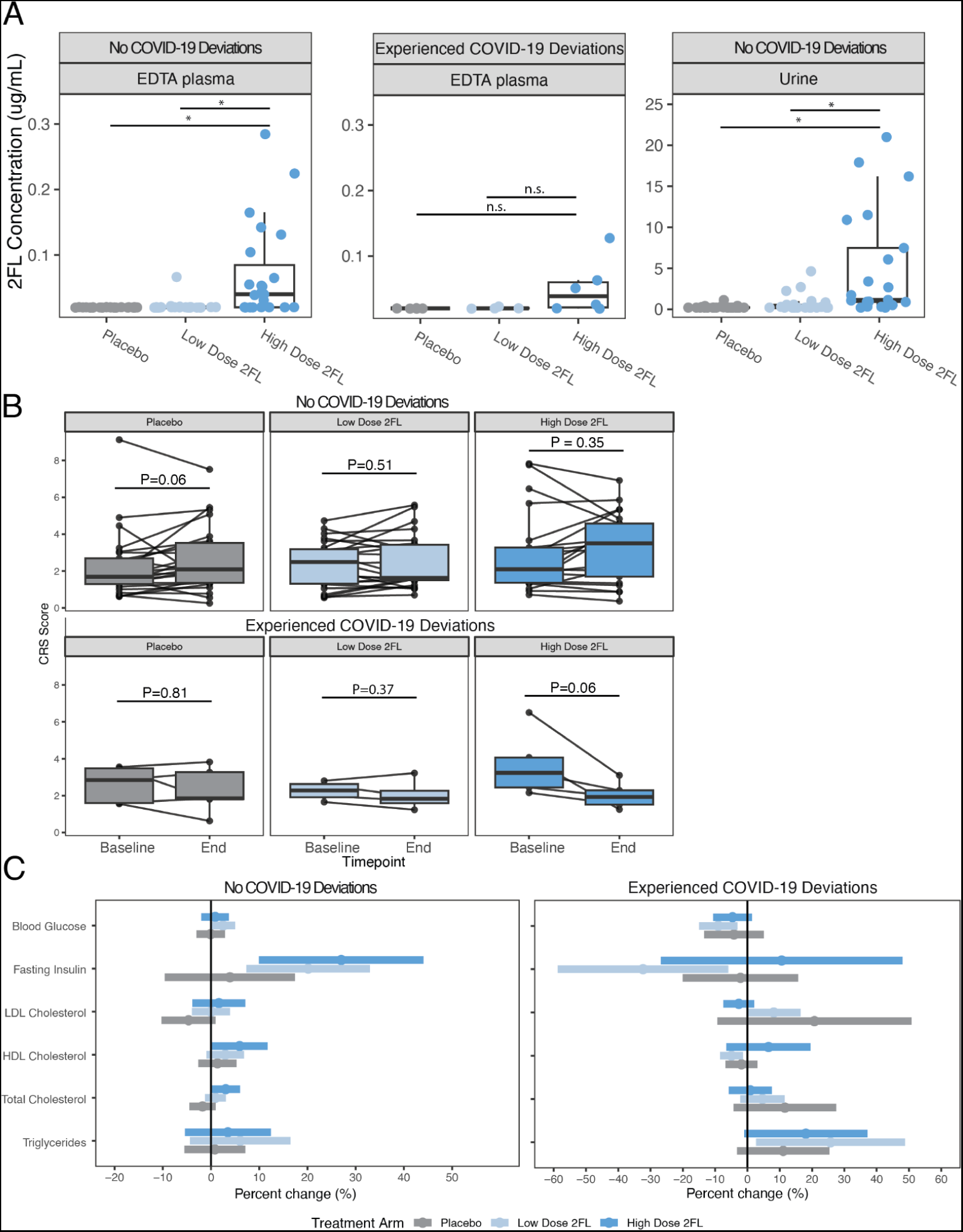
Sensitivity analysis of COVID-19 protocol deviations for plasma and urine 2’-FL quantification, Cytokine Response Score (CRS) and clinical markers. (A) Participants in the High Dose 2’-FL group who did not experience COVID-19 deviations had significantly higher 2’-FL compared to the Low Dose 2’-FL group and the Placebo group for both EDTA plasma (P = 0.004, P = 0.003, for Low Dose 2’-FL and Placebo comparisons respectively, Student’s t-test) and in urine. For the COVID-19 deviation group, there were no significant differences between the High Dose 2’-FL group and the Low Dose 2’-FL group (P = 0.13, Student’s t-test) and Placebo group (P = 0.12, Student’s t-test). However, as seen in the no deviation group, there was no 2’-FL detected in Placebo group participants. Note that participants who deviated from the protocol were unable to return urine samples at the end of intervention. **(B)** CRS analysis comparing baseline (Week 0) to end of intervention (Week 6) for the three treatment groups, separated by the subset of participants who did not experience a COVID-19 deviation (top) and the participants who did (bottom). None of the baseline-to-end differences are statistically significant (p-values listed in figure, all tests are paired Wilcoxon rank-sum tests). **(C)** Clinical panel results showing percent change from baseline to end of intervention for fasting glucose, fasting insulin, LDL cholesterol, HDL cholesterol, total cholesterol and triglycerides. Left panel is participants who did not experience COVID-19 protocol deviation, right panel is participants who did experience COVID-19 protocol deviations. Dots are group means for each treatment arm, bars are 95% confidence intervals. For the subset that did not experience COVID-19 protocol deviations we observed significant increases in fasting insulin in the High Dose 2’-FL group (95% CI: 9.9%, 44.1%) and the Low Dose 2’-FL group (95% CI: 7.3%, 33%), a significant increase in HDL cholesterol in the High Dose 2’-FL group (95% CI: 0.1%, 11.7%), and a significant increase in total cholesterol in the High Dose 2’-FL group (95% CI: 0.1%, 6%). The subset that did experience COVID-19 protocol deviations saw several significant changes in the Low Dose 2’-FL group: blood glucose (95% CI: −15%, −3.1%), fasting insulin (95% CI: −58.7%, −5.9%), HDL cholesterol (95% CI: −8.5%, −1.4%), and triglycerides (95% CI: 2.7%, 48.8%). However, it should be noted that there are only 4 individuals in this group and results are expected to be noisy with a sample size this large.

**Figure D2.**
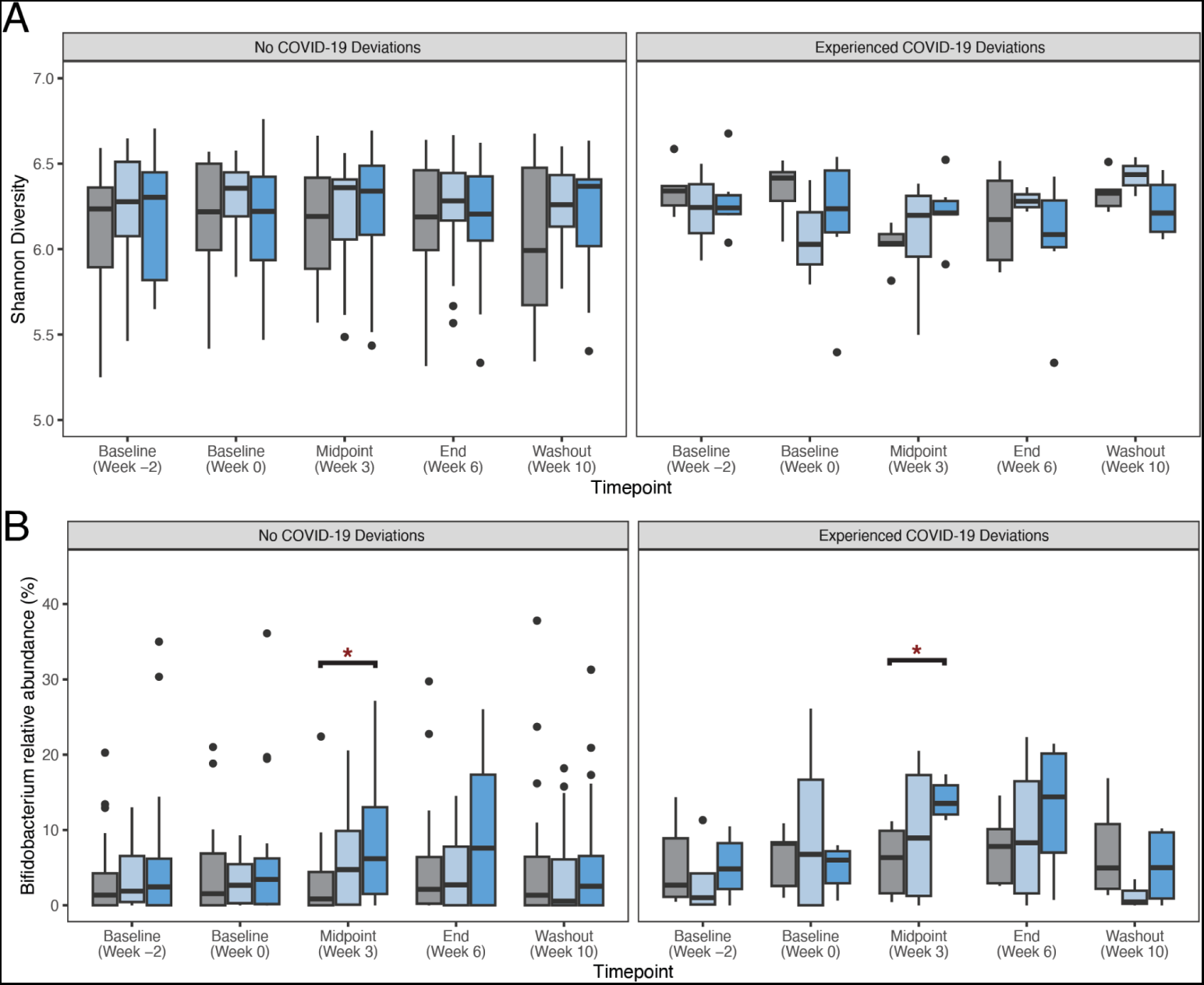
Sensitivity analysis of COVID-19 protocol deviations for alpha diversity and *Bifidobacterium* abundance. (A) Boxplots showing Shannon diversity for each treatment arm at each timepoint, broken out by the subset of participants that did not experience COVID-19 protocol deviations (left) and the subset that did experience COVID-19 protocol disruptions (right). None of the between-arm comparisons are significantly different at each timepoint for the subset that did not experience protocol deviations. None of the comparisons between the “No deviations” group and the “Experienced deviations” group for a given treatment arm and time point are significantly different. (B) Boxplots showing relative abundance of *Bifidobacterium* for each treatment arm at each time point, broken out by the subset of participants that did not experience COVID-19 protocol deviations (left) and the subset that did experience COVID-19 protocol disruptions (right). For both subsets of participants, *Bifidobacterium* abundance was significantly greater in the High Dose 2’-FL arm at Week 3 compared to the Placebo group (P = 0.01 for the no deviation group, P = 0.004 for the deviation group, Wilcoxon rank-sum tests). Bifidobacterium relative abundance was not significantly greater in the subset that experienced COVID-19 deviations compared to the group that did not experience COVID-19 deviations in the High Dose 2’-FL group at the Week 3 timepoint (P = 0.08, Wilcoxon rank-sum test).

### Supplemental Figures

**Figure S1.**
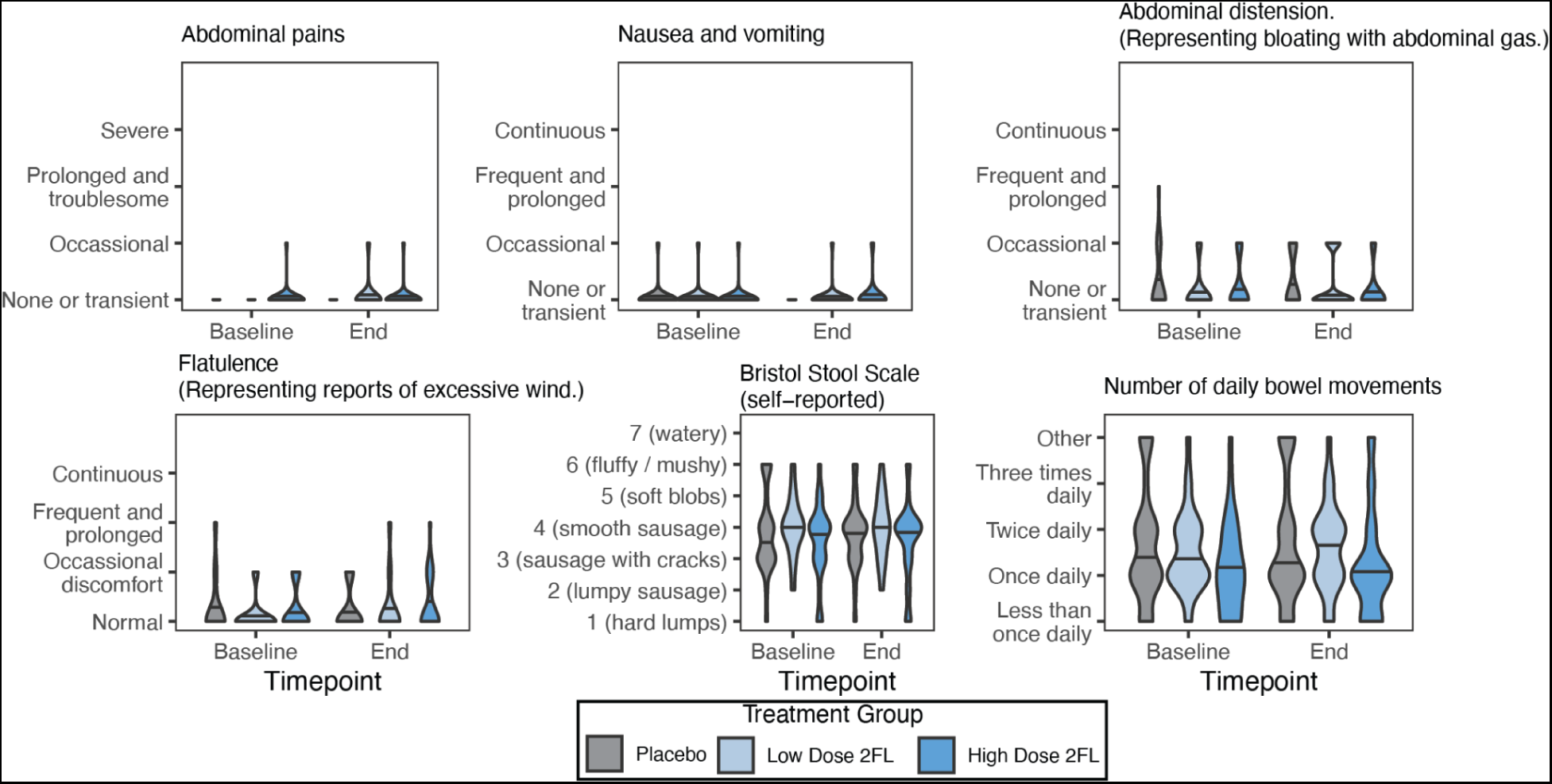
Results of gastrointestinal tolerance surveys showing that 2’-FL is well-tolerated. Self-reported results of gastrointestinal symptoms and bowel movements. There were no significant differences between treatment arms for any of the above categories and no adverse events reported.

**Fig S2.**
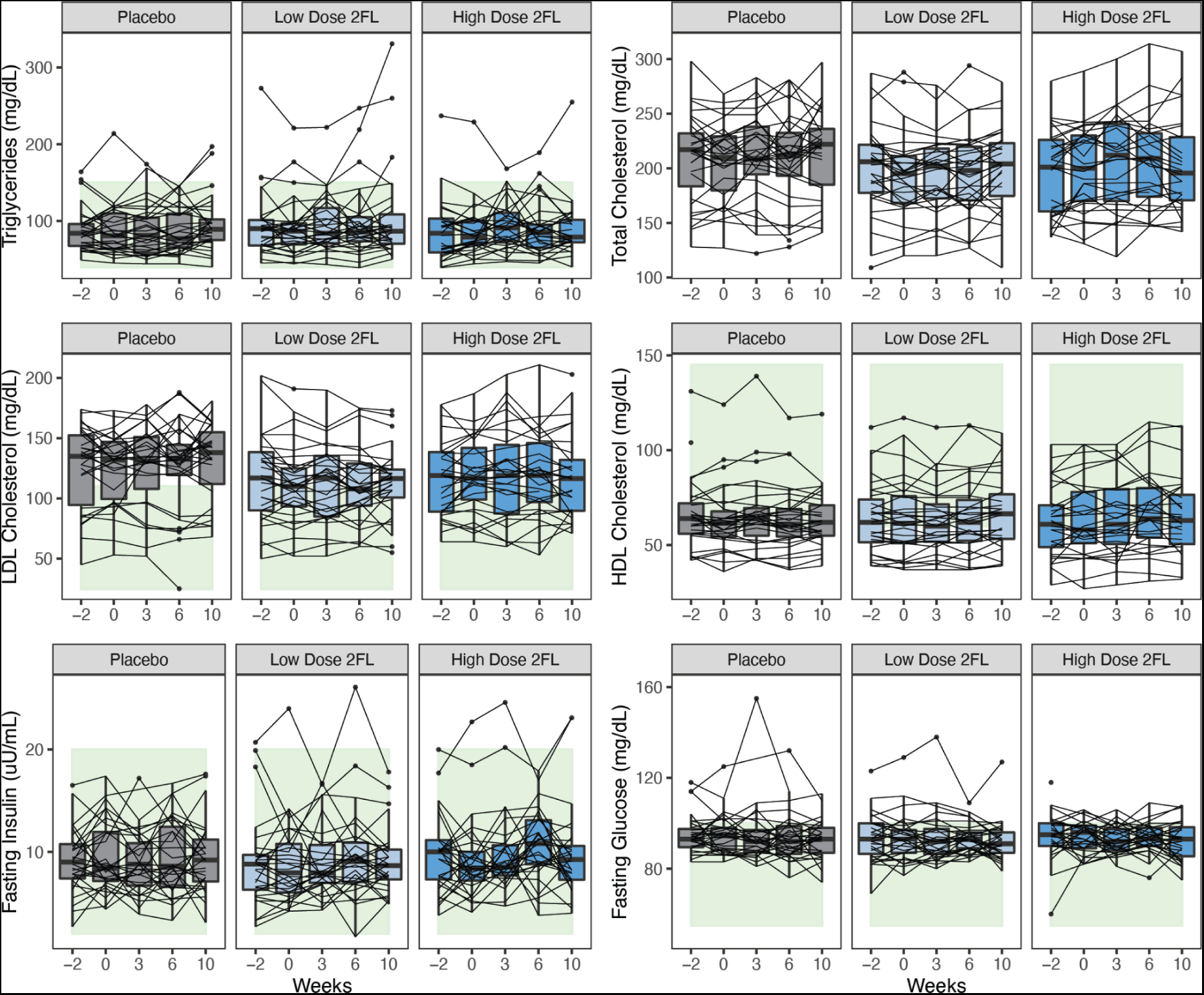
Raw values of results from standard clinical panels. Boxplots, separated by treatment arm, showing triglycerides, total cholesterol, LDL cholesterol, HDL cholesterol, fasting insulin and fasting glucose for each subject at 5 timepoints (Week −2, Week 0, Week 3, Week 6 and Week 10). Lines connect individual subjects over time. Green bands in background show standard healthy reference ranges.

**Figure S3.**
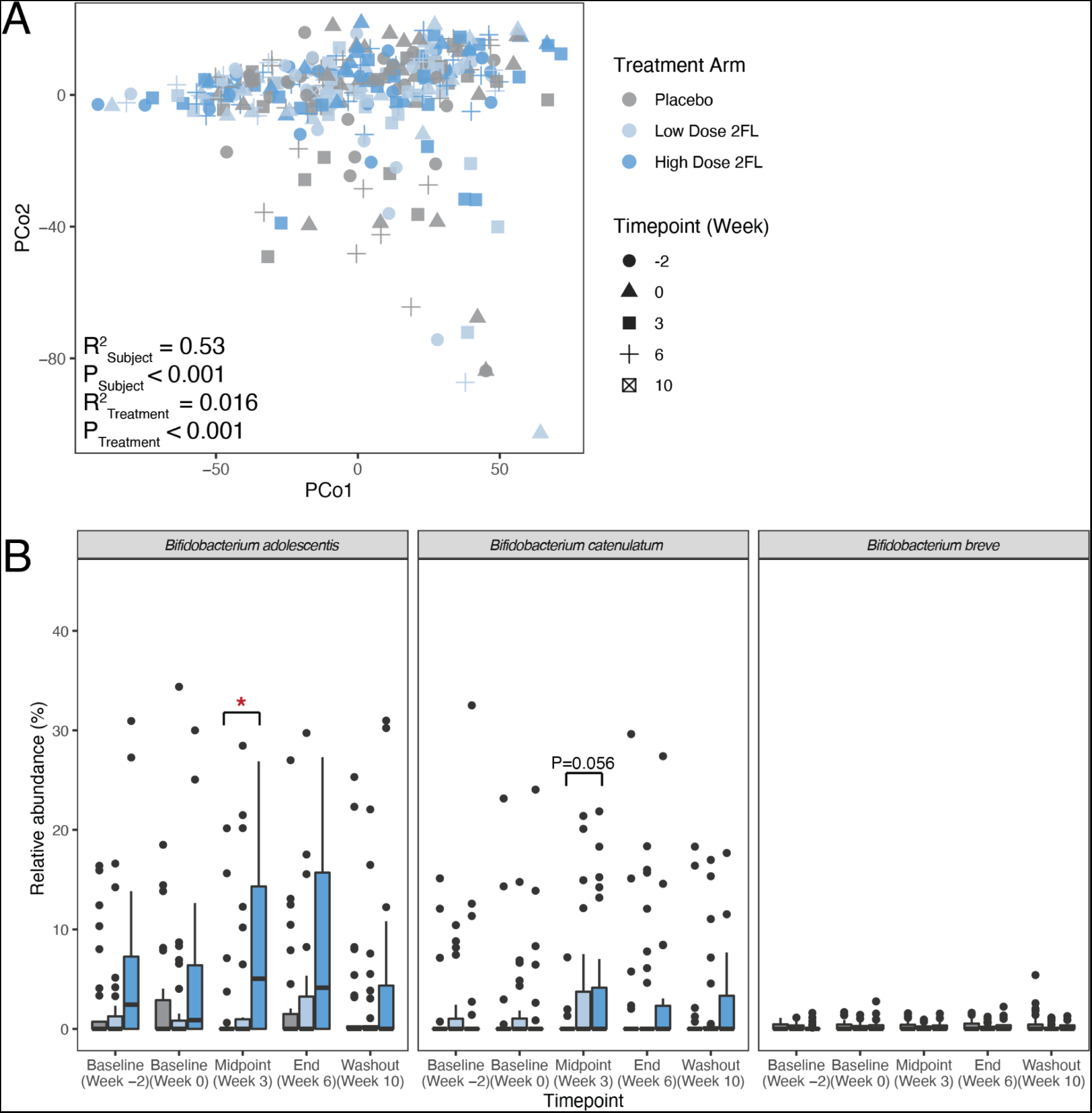
Metagenomics and species-specific analysis of the gut microbiota of RAMP participants. (A) A principal coordinate analysis plot of all stool samples across the three treatment arms based on euclidean distance of KEGG pathway abundance based on the Humann3 pipeline. Treatment arm explains a statistically significant portion of the variance (P<0.001, Adonis test). (B) Relative abundance of the 3 species of the genus *Bifidobacterium* that could be identified using SILVA 16S rRNA taxonomy. *Bifidobacterium adolescentis* showed greater relative abundance in the High Dose 2’-FL group compared to the Placebo group at the Week 3 timepoint (P=0.001, Wilcoxon rank-sum test). *Bifidobacterium catenulatum* also showed greater (but not statistically significant) relative abundance in the High Dose 2’-FL group compared to the Placebo group at the Week 3 timepoint (P=0.056, Wilcoxon rank-sum test).

**Figure S4.**
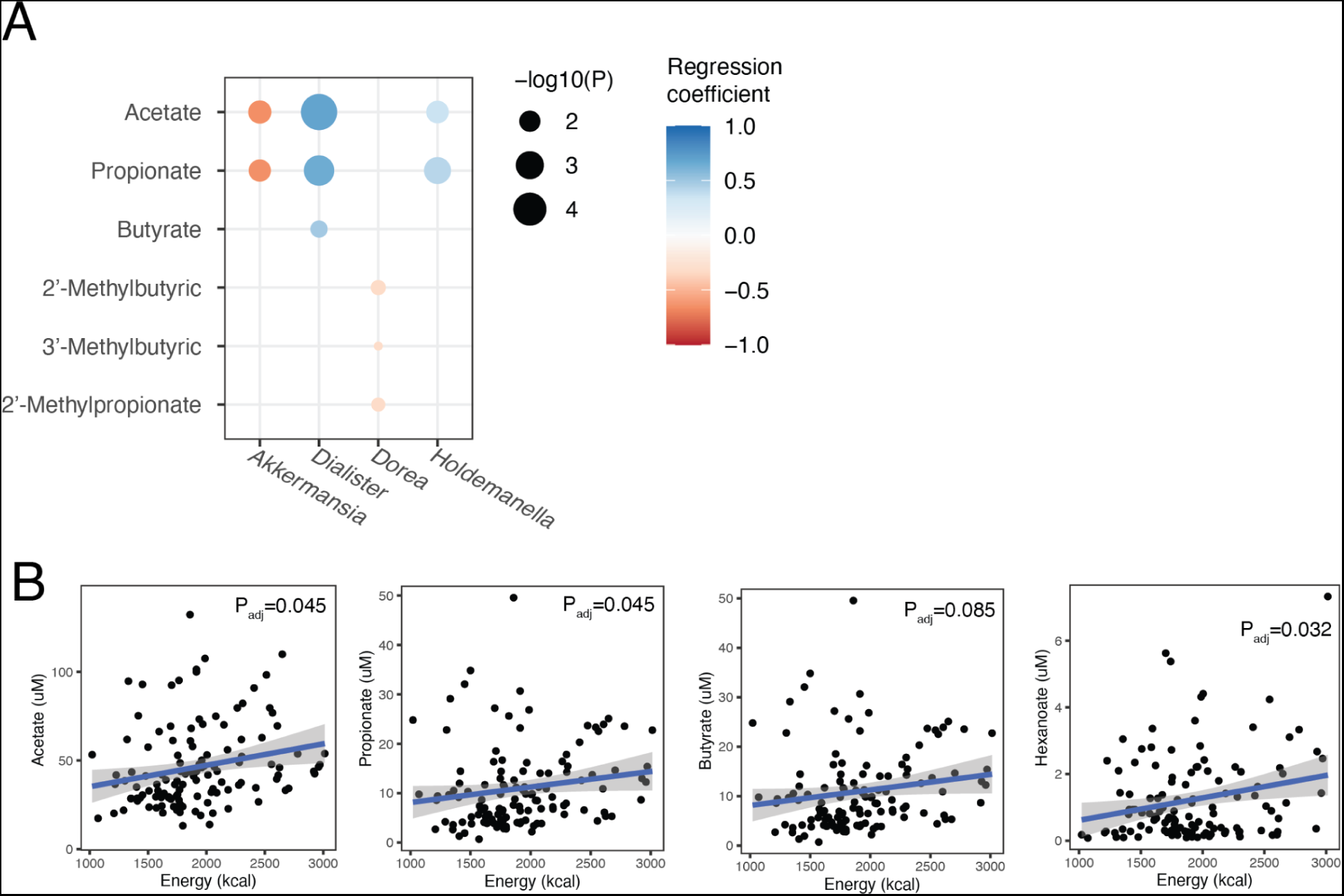
Significant associations between levels of stool SCFAs and microbial relative abundance and diet. (A) Statistically significant pairwise correlations between SCFAs and 4 genera found in the gut microbiota of RAMP participants. All P values were corrected using the Benjamini-Hochberg method. (B) Statistically significant pairwise correlations between SCFAs and daily energy consumption (in kilocalories) according to dietary logs from RAMP participants. All P values were corrected using the Benjamini-Hochberg method. Note that the adjusted P value for the butyrate association is not statistically significant. Participant identity was used as a random effect in these models to control for intra-participant correlation.

**Figure S5.**
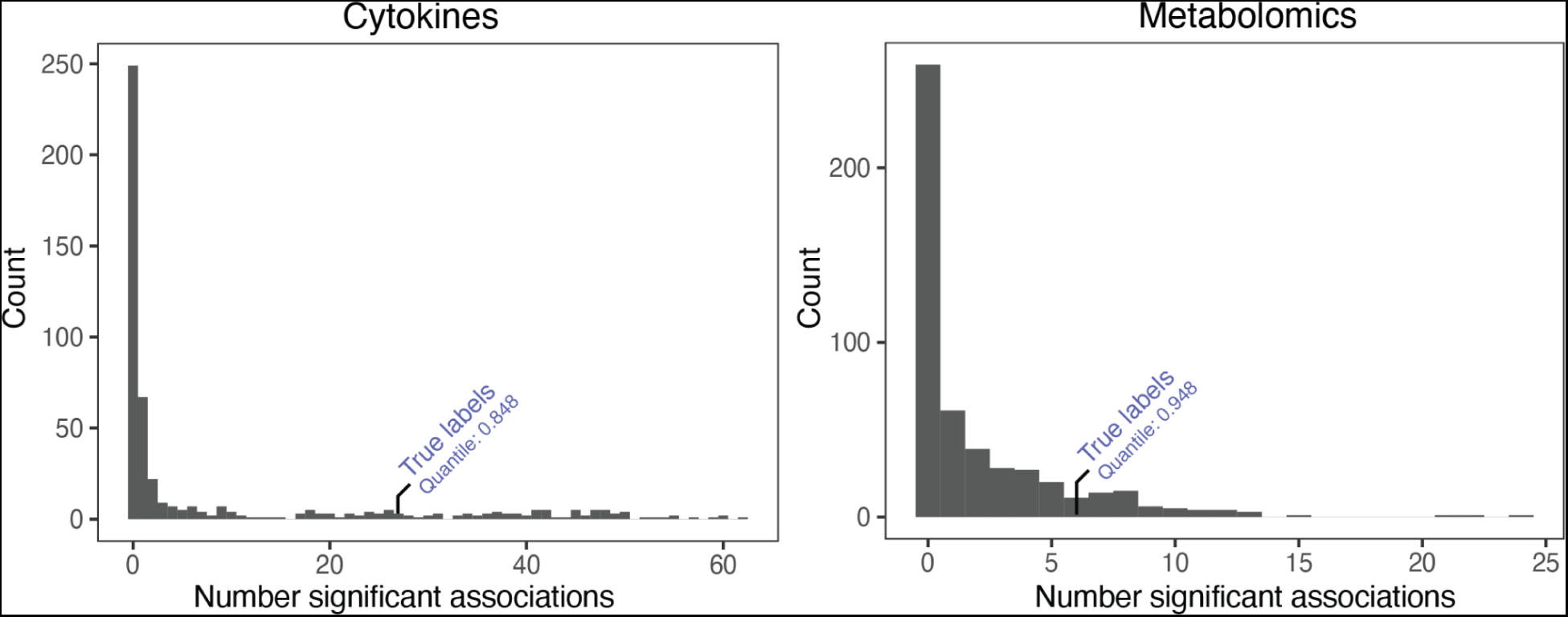
Simulated analyses of serum cytokine proteomics and serum metabolomics with permuted responder/nonresponder labels. Participants were assigned true responders/nonresponder labels on the basis of their change in *Bifidobacterium* abundance from Week 0 to Week 3. We identified 24 significantly enriched cytokines and 8 significantly enriched metabolites. We performed 500 simulations where we permuted the responder/nonresponder labels each time and determined the number of significantly enriched cytokines/metabolites each time. The histograms above show the number of significant associations recovered in each simulation. The number of significant cytokines recovered with the true labels was greater than 84.8% of all simulations. The number of significant metabolites with the true labels was greater than 94.8% of all simulations.

**Figure S6.**
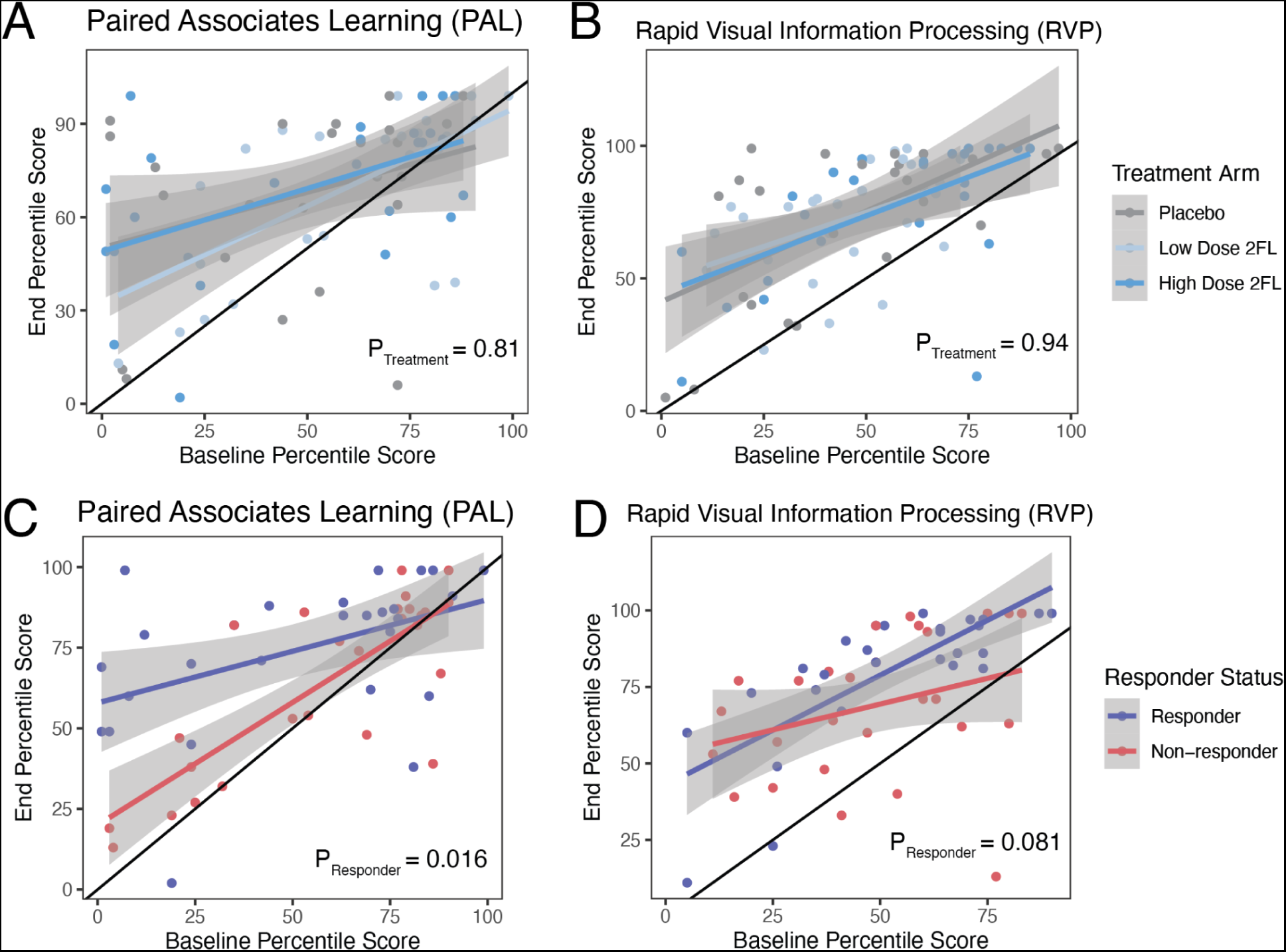
Results of cognitive surveys. (A) Scatter plot comparing baseline (x axis) and end of intervention (y axis) percentile scores on the Paired Associates Learning test from CANTAB, colored by treatment group. Analysis of covariance analysis showed no significant differences between treatment arms (P=0.81). Solid black line is 1:1 line. Individuals above 1:1 line performed better on the end of intervention test than their baseline test. (B) Scatter plot comparing baseline (x axis) and end of intervention (y axis) percentile scores on the Rapid Visual Information Processing test from CANTAB, colored by treatment group. Analysis of covariance analysis showed no significant differences between treatment arms (P=0.94). Solid black line is 1:1 line. (C) Scatter plot comparing baseline (x axis) and end of intervention (y axis) percentile scores on the Paired Associates Learning test from CANTAB, colored by responder status. Analysis of covariance analysis showed significant differences between responders and nonresponders (P=0.016). Solid black line is 1:1 line. (D) Scatter plot comparing baseline (x axis) and end of intervention (y axis) percentile scores on the Rapid Visual Information Processing test from CANTAB, colored by responder status. Analysis of covariance analysis showed no significant differences between responders and nonresponders (P=0.081). Solid black line is 1:1 line.

## STAR Methods

### RESOURCE AVAILABILITY

#### Lead contact

All information and requests for further resources should be directed to and will be fulfilled by the Lead Contacts, Justin Sonnenburg, and Christopher Gardner.

#### Materials availability

This study did not generate new unique reagents.

### EXPERIMENTAL MODEL AND SUBJECT DETAILS

#### Recruitment and selection of participants

Participants were recruited from the local community through online advertisement in different community groups as well as emails to past research participants that consented to being contacted for future studies. The current study assessed 335 participants for eligibility. They completed an online screening questionnaire and a clinic visit between March 2019 and December 2020. The primary inclusion criteria included age ≥ 60 years and general good health. Participants were excluded if they had a history of active uncontrolled inflammatory bowel disease (IBD) including ulcerative colitis, Crohn’s disease, or indeterminate colitis, irritable bowel syndrome (IBS) (moderate or severe), infectious gastroenteritis, colitis or gastritis, Clostridium difficile infection (recurrent) or Helicobacter pylori infection (untreated), malabsorption (such as celiac disease), major surgery of the GI tract, with the exception of cholecystectomy and appendectomy, in the past five years, or any major bowel resection at any time. Other exclusion criteria included a BMI ≥ 40, diabetes, renal disease, significant liver enzyme abnormality, smoking, a history of CVD, inflammatory disease, or malignant neoplasm. Consort flow diagram of participant recruitment shown in Figure 1B and demographics table shown in Table S1. 89 participants (49 female sex and gender identifying, 40 male sex and gender identifying) were used for full analysis with an average age of 67.3 ± 5.4 years. All study participants provided written informed consent. The objective of this study is to define the impact of a prebiotic supplement on microbiome, immune system, and metabolic status in older adults. The study was approved annually by the Stanford University Human Subjects Committee. Trial was registered at ClinicalTrials.gov, identifier: NCT03690999.

#### Specimen collection

Stool samples were collected at Weeks −2 (pre-baseline), 0 (baseline), 3 (midpoint of intervention), 6 (end of intervention), and 10 (washout). All stool samples were kept in participants’ home freezers (−20C) wrapped in ice packs, until they were transferred on ice to the research laboratory and stored at −80C. Blood samples were collected during research clinic visits at Weeks −2 (pre-baseline), 0 (baseline), 3 (midpoint of intervention), 6 (end of intervention), and 10 (washout). Blood for PBMC and whole blood aliquots were collected into heparinized tubes. Whole blood aliquots were incubated with Proteomic Stabilization Buffer (Smart tube, Fisher Scientific) for 12 minutes at room temperature and stored at −80C. PBMCs were isolated using Ficoll-Paque PLUS (Sigma-Aldrich), washed with PBS, frozen at −80C for 24 hours then moved to liquid nitrogen for longer storage. Blood for serum was collected into an SST-tiger top tube, spun at 1,200xg for 10 minutes, aliquoted, and stored at −80C. Blood for plasma was collected into an EDTA tube, spun at 1,200xg for 10 minutes, aliquoted, and stored at −80C.

### METHOD DETAILS

#### Randomization

Participants were randomized to the Placebo arm, the Low Dose 2’-FL arm or the High Dose 2’-FL arm. A simple randomization was done for the three groups using a random number generator (Excel), performed by a statistician not involved in the intervention or data collection.

#### Dietary logs and health questionnaires

Participants logged all their food and drink intake for 3 days (2 weekdays and 1 weekend) each week during the ramp phase and every other week for the rest of the study using the HealthWatch360 app. The dietitian reviewed the entries with participants to assess accuracy of entries and portions.

The following validated health surveys were used by participants: PROMIS v1.1 global health, PROMIS v1.0 - fatigue, WHO wellbeing index, PROMIS applied cognition short form, Perceived Stress Scale (Cohen et al., 1983), and the International Physical Activity Questionnaire (Craig et al., 2003).

#### 16S amplicon sequencing

DNA was extracted from stool using the MoBio PowerSoil kit according to the Earth Microbiome Project’s protocol (Gilbert et al., 2014) and amplified at the V4 region of the 16S ribosomal RNA (rRNA) subunit gene and 250 nucleotides (nt) Illumina sequencing reads were generated. There was an average of 60,192 reads per sample (standard deviation of 22,043) and samples with less than 2,000 reads were filtered out (2 samples out of 419 removed). There was an average of 40,621 reads per sample recovered (standard deviation of 16,022) after filtering, denoising, and removing chimeras. 16S rRNA gene amplicon sequencing data from both stool samples and fermented food samples were demultiplexed using the QIIME pipeline version 1.8 (Caporaso et al., 2010). Amplicon sequence variants (ASVs) were identified with a learned sequencing error correction model (DADA2 method) (Callahan et al., 2016), using the dada2 package in R. ASVs were assigned a taxonomy using the Silva database (version 138.1) ^48^. ɑ-diversity was quantified as the number of observed ASVs, Shannon diversity, or PD whole tree, in a rarefied sample using the phyloseq package in R (version 3.4.0). Data were rarefied to 6,727 reads per sample (lowest 5% of reads, 398 samples retained out of 419 total) also using the phyloseq package in R. Rarefied data were only used for ɑ-diversity measures. b-diversity was calculated using the ordinate function in the phyloseq package in R (version 3.4.0) for weighted and unweighted Unifrac. To determine if the treatment arms were significantly different at different timepoints, the samples were filtered to a given timepoint, beta diversity was calculated, and the analysis of variance using distance matrices was calculated with the adonis function (method = “bray”, permutations=5000) in the vegan package in R (version 2.5.6).

#### Metagenomic sequencing

DNA extraction for shotgun metagenome sequencing was done using the MoBio PowerSoil kit as described in the 16S amplicon sequencing methods. For library preparation, the Nextera Flex kit was used with a minimum of 10ng of DNA as input and 6 or 8 PCR cycles depending on input concentration. A 12 base pair dual-indexed barcode (CZ Biohub) was added to each sample and libraries were quantified using an Agilent Fragment Analyzer. They were further size-selected using AMPure XP beads (Beckman) targeted at a fragment length of 450bp (350bp size insert). DNA paired-end sequencing (2x146bp) was performed on a NovaSeq 6000 using S4 flow cells (CZ Biohub). The average sequencing depth for each sample was 14.2 million paired-end reads (or 4.05 giga base pairs). Data quality analysis was performed by demultiplexing raw sequencing reads and concatenating data for samples that required multiple sequencing runs for target depth before further analysis. BBtools suite (https://sourceforge.net/projects/bbmap/)) was used to process raw reads and mapped against the human genome (hg19) after trimming, with masks over regions broadly conserved in eukaryotes (http://seqanswers.com/forums/showthread.php?t=42552). Exact duplicate reads (subs = 0) were marked using clumpify and adapters and low-quality bases were trimmed using bbduk (trimq = 16, minlen = 55). Finally, reads were processed for sufficient quality using FastQC (https://www.bioinformatics.babraham.ac.uk/projects/fastqc/).

#### Microbiome functional profiling

The abundance of KEGG orthologous groups and modules were determined using the HUMAnN pipeline ^49^ with default parameters. The input to this pipeline were the processed reads described in the “Metagenomic sequencing” section above.

#### Stool short-chain fatty acids

Quantification of short-chain fatty acids was performed using an LC/MS-based technique described here ^50^, using a method adapted from ^51^.

#### Serum cytokines

Cytokine data were generated from serum samples submitted to Olink Proteomics for analysis using their provided inflammation panel assay of 92 analytes (Olink Inflammation). Data are presented as normalized protein expression values (NPX, Olink Proteomics arbitrary unit on log2 scale). We removed 29 proteins from the data set that were below the limit of detection in greater than 30% of samples.

#### PhosphoFlow

This assay was performed by the Human Immune Monitoring Center at Stanford University. Normal PBMCs were isolated by density-gradient centrifugation and cryopreserved. PBMC were thawed in warm media, washed twice and resuspended at 0.5×10^6 viable cells/mL. 200 ul of cells were plated per well in 96-well deep-well plates. Cytokine stock solutions were prepared at a concentration of 100 g/ml for each cytokine (IFN, IL-6, IL-7, IL-10, IL-2, or IL-21), or 10^6 U/ml for IFN. After resting for 2 hours at 37°C, cells were stimulated by adding 50 ul of cytokine stock solution and incubated at 37°C for 15 minutes. The PBMCs were then fixed with paraformaldeyde, permeabilized with methanol, and kept at −80°C overnight. The cells were washed with FACS buffer (PBS supplemented with 2% FBS and 0.1% soium azide), and stained with the following antibodies (all from BD Biosciences, San Jose, CA): CD3 Pacific Blue, CD4 PerCP-Cy5.5, CD20 PerCp-Cy5.5, CD33 PE-Cy7, CD45RA Qdot 605, pSTAT-1 AlexaFluor488, pSTAT-3 AlexaFluor647, pSTAT-5 PE. The samples were then washed and resuspended in FACS buffer. 100,000 cells per stimulation condition were collected using DIVA software on an LSRII flow cytometer (BD Biosciences). Data analysis was performed using FlowJo software (BD), by gating on live cells based on forward versus side scatter profiles, then on singlets using forward scatter area versus height, followed by cell subset-specific gating. The 90th percentile fluorescence for each pSTAT marker on each cell population was reported.

#### Untargeted serum metabolomics

Serum samples were extracted in LC-MS grade methanol (4:1 v/v). These serum samples were identical to the samples used for serum cytokine analysis detailed above. Protein precipitation was conducted by incubating the samples for 5 minutes at room temperature and centrifugation at 5,000xg for 10 minutes. Sample supernatants were then transferred, evaporated, and reconstituted in an internal standard mix (50% Methanol). Metabolite samples were analyzed on a LC-MS qTOF instrument using reverse phase C18 positive mode as described ^52,53^. Compound annotation was carried out using the MSDIAL software (Tsugawa et al., 2015) and an authentic standard reference library. To quantify metabolite levels, area under the curve for each annotated metabolite was normalized using the sum of internal standards in each sample.

### QUANTIFICATION AND STATISTICAL ANALYSIS

#### Location of Statistical Details in the Text

Results of each experiment can be found in the results and figure legends. Significant values of statistical tests are also indicated by asterisks in the figures.

#### Statistical Analysis of Primary Outcome

The primary outcome as listed on ClinicalTrials.gov was the change in Cytokine Response Score within each arm from baseline (week 0) to end of intervention (week 6). CRS was calculated using the method described in (Shen-Orr et al., 2016). Significant changes were evaluated using Wilcoxon rank-sum tests.

#### Quantification of alpha and beta diversity measures

Alpha and beta-diversity measures were calculated using the phyloseq package (McMurdie and Holmes, 2013) in R. Alpha diversity was calculated at the ASV level. Bray-Curtis dissimilarity was computed at each timepoint and for each subject using the vegdist function in the R vegan package (Dixon, 2003).

#### Testing for significant changes in microbe and host features during the intervention

For each data type, we constructed linear mixed effect models with Maaslin2 ^54^. Each model was constructed using the treatment group as a fixed effect and the interaction between the treatment group and the intervention phase (“Baseline” or “Intervention”). Subject identity was used as a random effect. Features were deemed to vary significantly by treatment arm if the interaction term had a Benjamini-Hochberg-corrected p-value (q-value) less than 0.05. For 16S data, we used a centered-log ratio transformation on the input data matrix. For the metabolomics and proteomics data, we used a log2 transformation on the input data matrix.

#### Centering and Scaling Data

In analyses comparing different data types to each other, parameters were centered and scaled by column for the purposes of data regularization. All methods describing data as centered and scaled were done using the scale function in base R (scale function with parameters: center = TRUE, scale = TRUE).

#### Identifying significant correlations in serum proteomics and serum metabolomics data sets

First, centered and scaled each feature from the serum proteomics (Olink), serum untargeted metabolomics and clinical panel data sets. We imputed missing values in the data using the “impute.knn” function from the R package impute (v1.7). Then, we determined the change in Z-score from baseline-to-end for each participant. We then calculated an all-versus-all Pearson correlation for all features using the function “rcorr” from the R package Hmisc (v4.7). We calculated the Benjamini-Hochberg corrected p-value for each correlation and filtered the correlation matrix to contain only correlations with adjusted p-values < 0.05. We then generated a correlation network from this filtered correlation matrix using the R package igraph (v1.3.4).

### ADDITIONAL RESOURCES

Clinical trial registry #NCT03690999: https://www.clinicaltrials.gov/ct2/show/NCT03690999

